# Effect of daily periodic human movement on dengue dynamics: the case of the 2010 outbreak in Hermosillo, Mexico

**DOI:** 10.1101/2020.12.17.20248375

**Authors:** Mayra R. Tocto-Erazo, Daniel Olmos-Liceaga, José A. Montoya

## Abstract

The human movement plays an important rol in the spread of infectious diseases. On an urban scale, people move daily to workplaces, schools, among others. Here, we are interested in exploring the effect of the daily local stay on the variations of some characteristics of dengue dynamics such as the transmission rates and local basic reproductive numbers. For this, we use a two-patch mathematical model that explicitly considers that daily mobility of people and real data from the 2010 dengue outbreak in Hermosillo, Mexico. Based on a preliminary cluster analysis, we divide the city into two regions, the south and north sides, which determine each patch of the model. We use a Bayesian approach to estimate the transmission rates and local basic reproductive numbers of some urban mobility scenarios where residents of each patch spend daily the 100% (no human movement between patches), 75% and 50% of their day at their place of residence. For the north side, estimates of transmission rates do not vary and it is more likely that the local basic reproductive number to be greater than one for all three different scenarios. On the contrary, tranmission rates of the south side have more weight in lower values when consider the human movement between patches compared to the uncoupled case. In fact, local basic reproductive numbers less than 1 are not negligible for the south side. If information about commuting is known, this work might be useful to obtain better estimates of some contagion local properties of a patch, such as the basic reproductive number.

## 1. Introduction

An important factor in the spread of infectious diseases as dengue is the human movement [1]. Due to this fact, dengue can be expanded from endemic to non-endemic places [2]. It has been suggested that infectious diseases may persist in a region where transmission rates are very low due to interaction with people from other areas with high transmission rates [3]. On an urban scale, daily movement occurs motivated by commuting people to workplaces, schools, commerce, among others [4].

Human mobility has been included in the latest generation of models in epidemiology using two main approaches: agent-based modeling and metapopulations [5]. Metapopulation models divide the population into interacting population groups defined by spatial or demographic information [6]. This mathematical modeling approach, based on ordinary differential equations, has been used to theoretically evaluate the effect of human mobility on the dynamics of infectious diseases in heterogeneous regions connected by the mobility [3, 7–11]. The consequences of mobility between cities have been analyzed in [12–14], whereas between an urban and suburban area in [15–17], or between areas within the same city in [18]. There are some efforts to use real data to validate the effect of human movement but there are few studies yet [14, 16, 18]. Authors in [14, 18] have concluded that infectious diseases could spread in disease-free areas or with local basic reproductive number less than one due to the human movement. In particular, authors in [14] study different factors for the transmission of dengue disease in a Chinese province using a residence time approach. They analyze the human movement between seven regions and estimate model parameters for each patch based on a fixed residence time matrix and explore some hypothetical scenarios by reducing the values of such matrix.

Similar to [14], we are interested in studying the daily human mobility between two regions from an urban area, to explore how the transmission rates and the local basic reproductive numbers may vary depending on the time period of daily local stay of a population within their own region. For this, we use a two-patch mathematical model under a little explored approach [15, 19], and data from the 2010 dengue outbreak in Hermosillo, Mexico. We use the ideas of a previous work [19] and applied them to a scenario where the commutation between patches emerges naturally. To define each of two patch of the model, we divide the city of Hermosillo into two regions, which was derived from a preliminary cluster analysis. We use a Bayesian approach to obtain estimates of some parameters of the model and compare mobility scenarios.

This work is divided into the following sections. The description of the model used, the data and the inference method are given in Section 2. Then, in Section 3 we show the estimation results. Finally, the conclusions and discussions on our results are presented in Section 4.

## 2. Methods

### 2.1. Mathematical model

We consider a previous two-patch model without vital dynamics in humans and with human daily movement, where movement takes place at periodic discrete times [19]. Here the interval [*t*_*k*_, *t*_*k*+1_) represent the *k*th day and is divided into two time periods: low-activity period [*t*_*k*_, *t*_*k*_ +*T*_*l*_) and high-activity period [*t*_*k*_ +*T*_*l*_, *t*_*k*+1_), where *T*_*l*_ represents the fraction of the *k*th day of low activity and *T*_*l*_ *∈* (0, 1). The daily dynamics between the periods of low-activity and high-activity are as follows. At the beginning of high-activity periods, people move to the other patch to carry out their daily activities. Then, at the end of high-activity periods people return to their residence patch and stay there during the low-activity periods. Thus, the following equations represent the dynamics of the populations for the low-activity period [*t*_*k*_, *t*_*k*_ + *T*_*l*_):

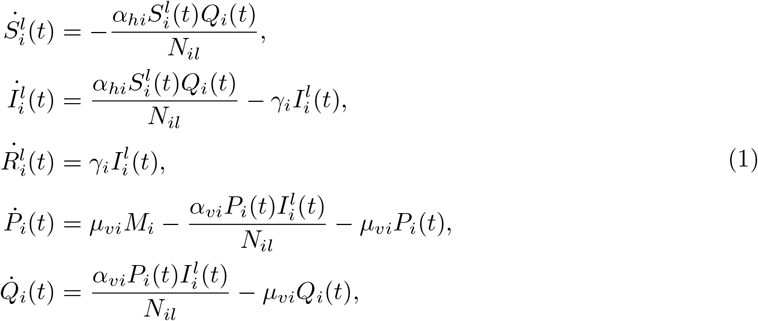

where *N*_*il*_ := *N*_*i*_ and *i* = 1, 2. For the high-activity period [*t*_*k*_ + *T*_*l*_, *t*_*k*+1_), the set of equations become:

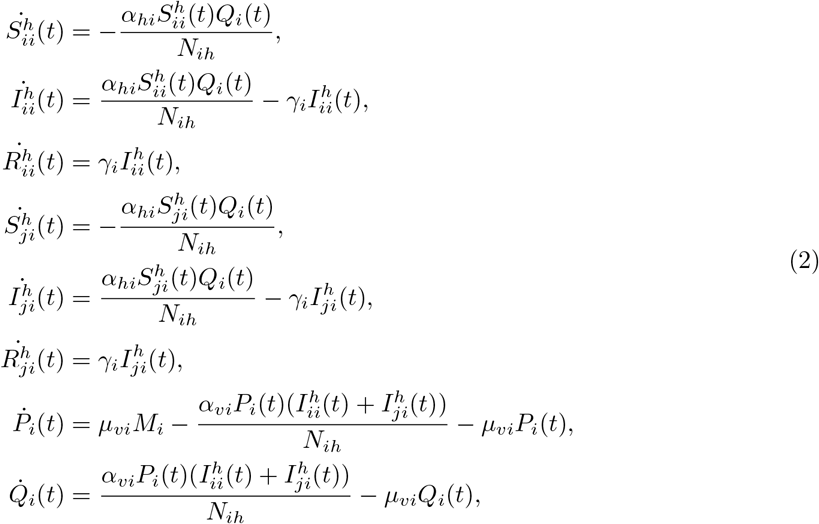

where *N*_*ih*_ := (1 *− α*_*i*_)*N*_*i*_ + *α*_*j*_*N*_*j*_, and *i, j* = 1, 2, *i /*= *j*. All model parameters and meaning of the state variables are defined in Table 1.

**Table 1:** Parameter definition of model (1)-(2).

| Variable or parameter | Meaning |
| --- | --- |
| $N_i$ | Total population from patch $i$. |
| $N_{ih}$ | Total population in patch $i$ during the high-activity period. |
| $M_i$ | Total population of mosquitoes from patch $i$. |
| $S_i^l$ | Susceptible residents from patch $i$. |
| $I_i^l$ | Infected residents from patch $i$. |
| $R_i^l$ | Recovered residents from patch $i$. |
| $S_{ii}^h$ | Susceptible residents from patch $i$ who do not commute to another patch during the low-activity period. |
| $I_{ii}^h$ | Infected residents from patch $i$ who do not commute to another patch during the low-activity period. |
| $R_{ii}^h$ | Recovered residents from patch $i$ who do not commute to another patch during the low-activity period. |
| $S_{ji}^h$ | Susceptible residents from patch $j$ who commute to patch $i$ during the high-activity period. |
| $I_{ji}^h$ | Infected residents from patch $j$ who commute to patch $i$ during the high-activity period. |
| $R_{ji}^h$ | Recovered residents from patch $j$ who commute to patch $i$ during the high-activity period. |
| $\alpha_{hi}$ | Transmission rate from mosquito to human in patch $i$. |
| $\alpha_{vi}$ | Transmission rate from human to mosquito in patch $i$. |
| $1/\gamma_i$ | Average recovery time of humans in patch $i$. |
| $1/\mu_{vi}$ | Average life time of mosquitoes in patch $i$. |
| $\alpha_i$ | Proportion of humans from patch $i$ who move to patch $j$ at time $t_k + T_l$. |

We assume that the infected classes both 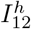 and 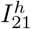, who move between patches, represent only individuals with mild or no symptoms. On the other hand, uncoupled case (*T*_*l*_ = 1) is obtained considering *α*_1_ = *α*_2_ = 0 in model (1)-(2). That is, system (1)-(2) is reduced to the dynamics of a vector-host model as in (1) for all time *t*.

Using the next generation matrix approach as in [20], the basic reproductive number (*R*_0*i*_) of uncoupled case is given by

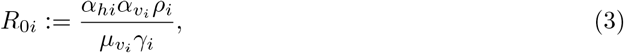

where *ρ*_*i*_ = *M*_*i*_*/N*_*i*_. Given the complexity of the model, the basic reproductive number for model (1)-(2) was not found. However, *R*_01_ and *R*_02_ can give us an approximation of a local indicator of the severity of the disease for both the uncoupled and the coupled cases.

### 2.2. Data

Hermosillo is a city located in the north of Mexico, with a total population of 715061 inhabitants according to the 2010 Census data provided by the National Institute of Statistics, Geography and Informatics from Mexico (INEGI). Based on preliminary cluster analysis and using a Geographic Information System (GIS), we divide the city into two areas: north and south side (Figure 1). The north side has 374102 inhabitants and the south side 340959. On the south side are located the offices of the municipal and state government, the city center, the largest university, industrial parks, among others. Thus, we consider that the flow from north to south of the city is greater than from south to north.

**Figure 1:**
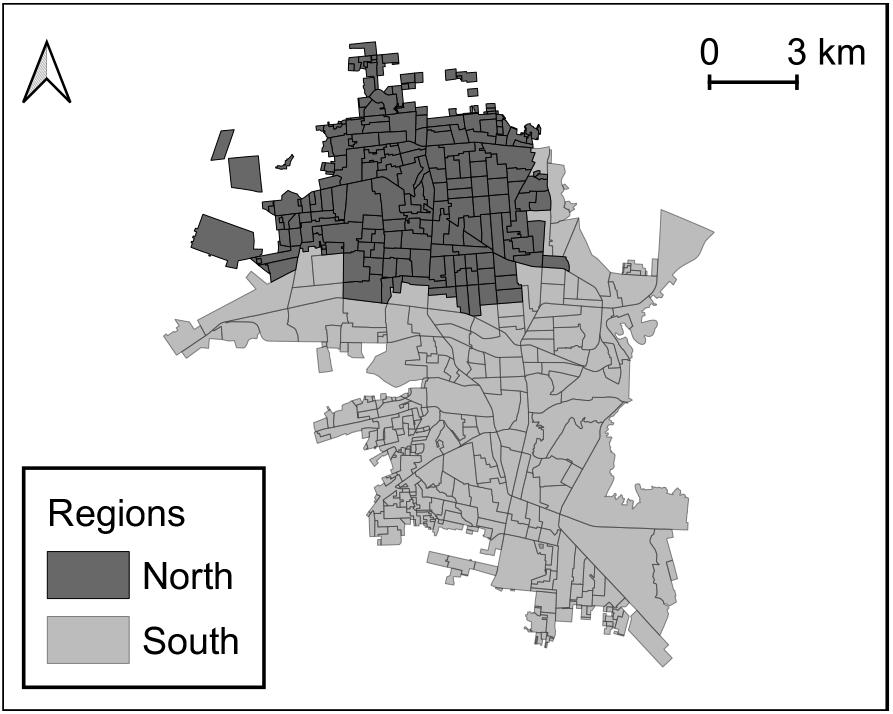
Division of Hermosillo in two regions: north (dark gray) and south (light gray).

According to data provided by the Health Ministry of the State of Sonora, 2139 dengue cases were located on the north along 52 epidemiological weeks resulting in a rate of 57.17cases*/*10000 inhabitants, and 590 dengue cases on the south with a rate of 17.3cases*/*10000 inhabitants during 2010. In this study, we use the weekly incidence from epidemiological week 33 to 40 for both regions north and south from Hermosillo.

### 2.3. Parameter estimation

For parameter estimation purposes, we add a class *C*_*i*_ to system (1)-(2), which represents the accumulated number of reported infected residents from patch *i*, and is given by

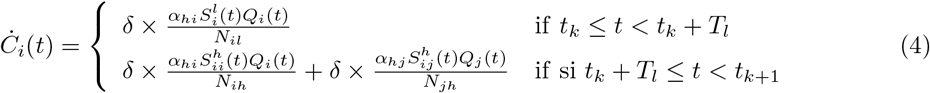

where, *δ* represents the proportion of infected individuals that were confirmed by the surveillance system of the state government of Sonora.

We establish the following movement scenario: *α*_1_ = 0.45 and *α*_2_ = 0.1. The first is determined based on the economically active population of the north side according to the 2010 Census data from Mexico, and the second is an assumed value considering that the daily flow of people is less from south to north. Based on previous studies, we assume that the average lifetime of a mosquito (1*/µ*_*vi*_) is 2 weeks [21], and the number of mosquitoes per person (*ρ*) is 2 [22]. Given a *T*_*l*_ fixed value, we use the data to estimate the remaining seven parameters (*α*_*h*1_, *γ*_1_, *α*_*v*2_, *α*_*h*2_, *γ*_2_, *α*_*v*2_, and *δ*) by Bayesian inference approach.

For computational purposes, we take logarithmic transformations as

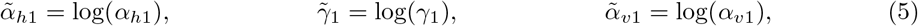

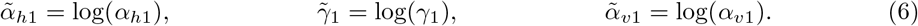

Thus, we assign a priori beta distribution for *δ* with shape parameters *α* = 5 and *β* = 50, and priori normal distributions for all six parameters given in (5)-(6) [23]. The distribution of 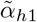 and 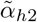 has a mean equal to log(0.3) and a standard deviation of 0.4. The distribution of 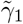 and 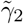 has a mean equal to log(0.22) and a standard deviation of 0.1. The distribution of 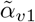 and 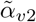 has a mean equal to log(0.4) and a standard deviation of 0.3. The distribution of 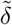 has a mean equal to log(0.08) and a standard deviation of 0.15. To establish the mean and standard deviation of the normal distributions for parameters (5)-(6), we considered the ranges given in [21]. Therefore, the prior joint density function of 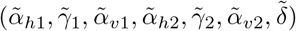 is given by

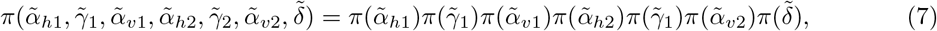

where *π*(*•*) is the normal density function of each parameter defined above.

Since model (1)-(2) is in a time-scale of days and starts at *t* = 0, we define 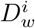 to represent the number of new infectious cases of dengue at *w*th week (*w* = 33, …, 40) in patch *i*, which is given by

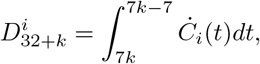

where *k* = 1, 2, …, 8, and *C*_*i*_(*j*) defined as (4). We define patch 1 as the north area and patch 2 as the south area. Thus, we consider that the new weekly cases at week *w* from patch 1 and patch 2 follow a Poisson distribution with a mean 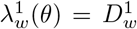 and 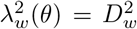, respectively, where 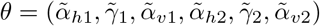. Thus, the sampling distribution is given by

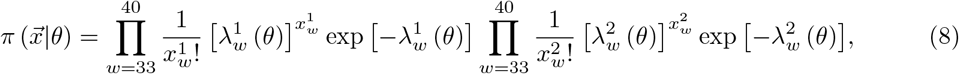

where 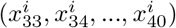 is the observed data from patch *i*, and 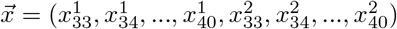.

Therefore, the posterior distribution 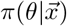 is given by

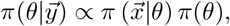

where *π*(*θ*) and 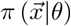 are given in (7) and (8), respectively.

To obtain the estimated probability density of the model parameters, we used a MCMC method based on the Metropolis-Hasting algorithm [24]. We run the algorithm for 200000 iterations, and use the last 50000 to generate the posterior densities of the parameters and the posterior predictive distributions to check the fit. The initial conditions assumed are as follows: 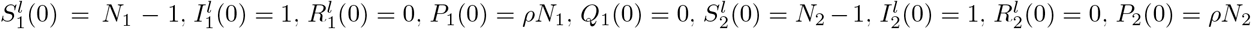 and *Q*_2_(0) = 0, where *N*_1_ = 374102 and *N*_2_ = 340959. To generate the posterior density of *R*_01_ and *R*_02_, we replace each sample of (*α*_*h*1_, *γ*_1_, *α*_*v*1_, *α*_*h*2_, *γ*_2_, *α*_*v*2_, *δ*) in equation (3). The code used to calculate the posterior predictive distribution is available online at https://github.com/MayraTocto/DailyHumanMobility_DengueOutbreak.

## 3. Results

The results are based on three scenarios: *T*_*l*_ = 1, *T*_*l*_ = 0.75 and *T*_*l*_ = 0.5. Case *T*_*l*_ = 1 represents that there is no flow of people between the north and south regions. Cases *T*_*l*_ = 0.75 and *T*_*l*_ = 0.5 indicate that the residents from each patch spend 75% and 50% of their day at their residence place, respectively. To check the fit of the model to the data, we construct 95% predictive intervals of the posterior predictive distributions using the 0.025 and 0.975 quantiles for the north and south sides, as shown in Figure 2. This figure shows that the three scenarios are plausible. In the next lines, we will see that there are differences in the estimates of the model parameters and, consequently, the estimated local basic reproductive numbers may drastically change.

**Figure 2:**
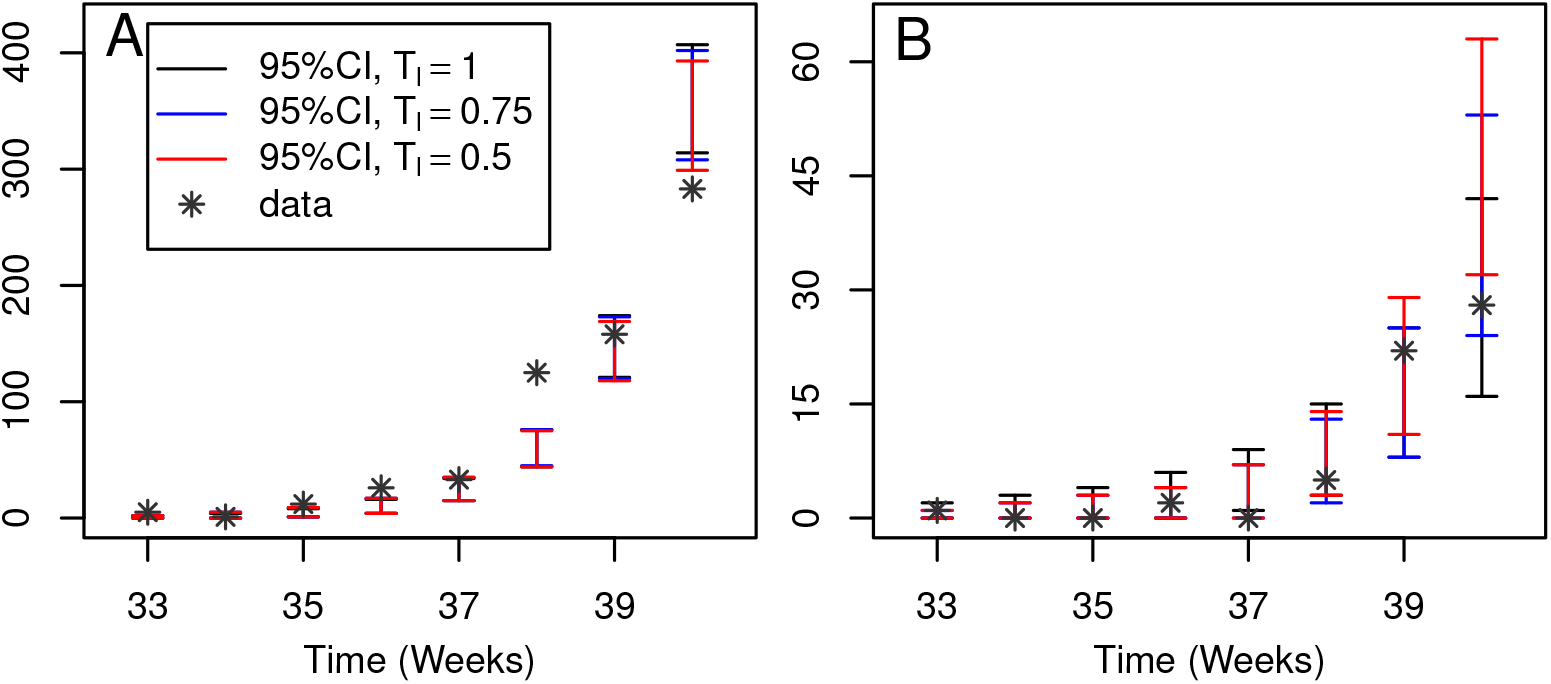
The 95% predictive intervals for the estimated posterior predictive distributions from the north side (A) and south side (B) versus data from dengue cases in Hermosillo.

Figure 3 shows the posterior densities of transmission rates for the north and south regions. The range of most likely values for estimated local transmission rates from the north side (*α*_*h*1_ and *α*_*v*1_) coincides for *T*_*l*_ = 1, *T*_*l*_ = 0.75 and *T*_*l*_ = 0.5 (Figure 3A and 3B). Based on our mobility scenarios, this fact could indicate that the daily mobility of people does not affect the disease dynamics on the north side of the city. On the contrary, the posterior density for local transmission rates (*α*_*h*2_ and *α*_*v*2_) from the south region, when *T*_*l*_ *<* 1, have more weight in lower values compared to the case *T*_*l*_ = 1 (Figure 3C and 3D). That is, it would be more likely to obtain smaller values for the transmission rates on the south side when *T*_*l*_ goes down from 1 to 0.5, and, therefore, local contagions decrease. In this case, according the *T*_*l*_ value (*T*_*l*_ *<* 1), infected individuals from the south side who get the dengue virus during their visit to the north side is more credible than local contagions. Thus, the results may suggest that some dengue cases from the south side may have emerged by the interaction between a susceptible individual from the south side that gets bitten by a infected mosquito from the north of the city during his visit. The median and 95% confidence intervals of the posterior distributions for *γ*_1_, *γ*_2_, and *δ* are given in Table 2.

**Figure 3:**
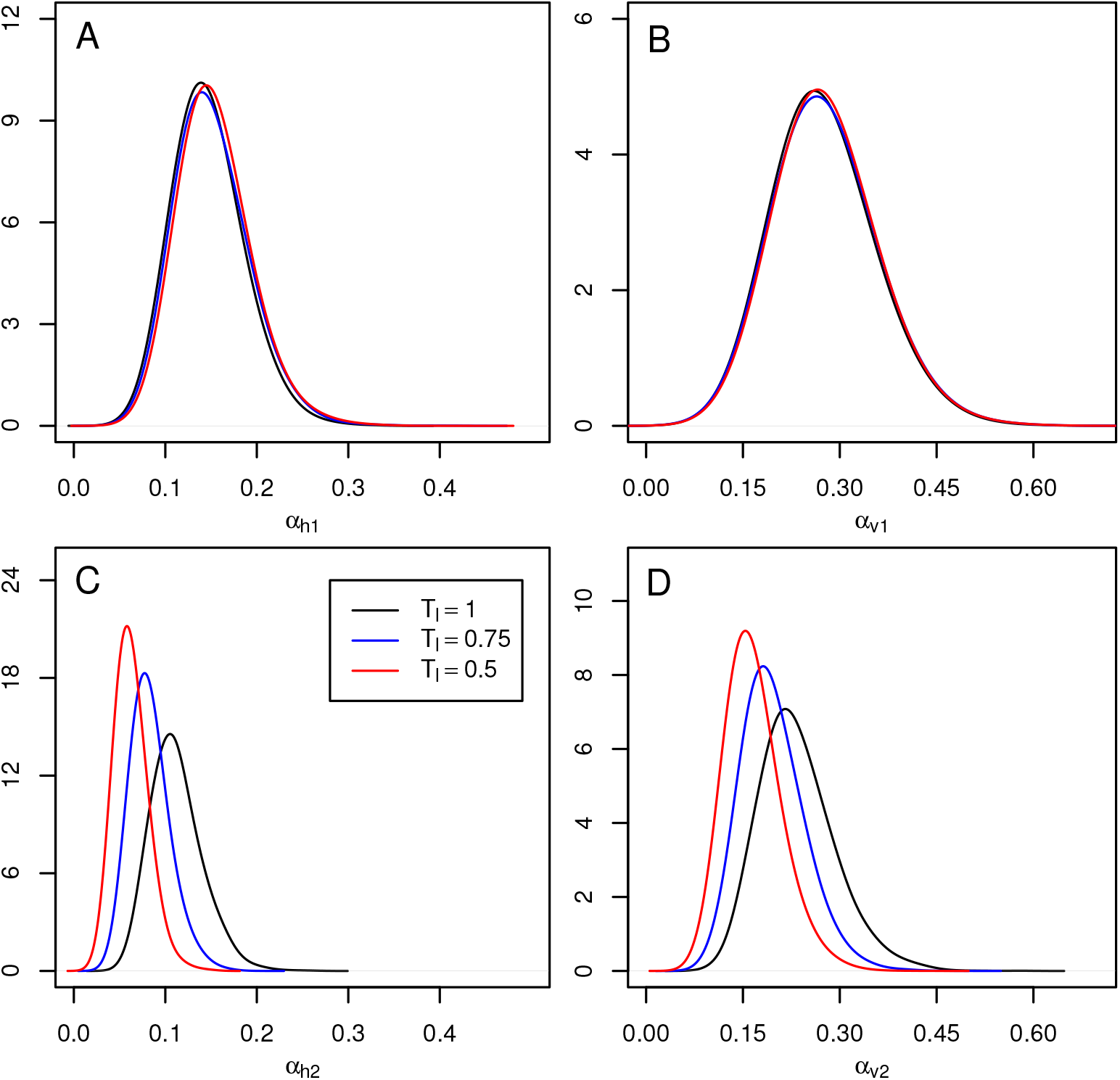
The posterior density of parameters from north patch (A and B) and south patch (C and D).

**Table 2:** Parameter estimation of *γ*_1_, *γ*_2_, and *δ* parameters.

| Parameter | Median (95% CI)<br>for $T_l = 1$ | Median (95% CI)<br>for $T_l = 0.75$ | Median (95% CI)<br>for $T_l = 0.5$ |
| --- | --- | --- | --- |
| $\gamma_1$ | 0.250 ([0.206, 0.305]) | 0.252 ([0.207, 0.307]) | 0.253 ([0.208, 0.309]) |
| $\gamma_2$ | 0.230 ([0.189, 0.280]) | 0.231 ([0.192, 0.283]) | 0.233 ([0.192, 0.284]) |
| $\delta$ | 0.429 ([0.337, 0.526]) | 0.436 ([0.344, 0.530]) | 0.436 ([0.342, 0.532]) |

Based on the samples obtained for the parameters and replacing them in expression (3), we obtain the estimated density of local basic reproductive numbers (for *T*_*l*_ = 1, *T*_*l*_ = 0.75 and *T*_*l*_ = 0.5). As we have already mentioned, for *T*_*l*_ *<* 1, the local basic reproductive number are used as local indicators of the disease, that is, these values would tell us how serious the disease could be locally. Figure 4 shows the posterior densities of *R*_01_ and *R*_02_. From Figure 4A, we have that *R*_01_ is more likely to take values greater than 1 for all three cases. However, if we consider *T*_*l*_ *<* 0.75, we have that the most probable values for *R*_02_ are smaller compared to the case *T*_*l*_ = 1 as we can see in Figure 4B. Furthermore, if we consider that *T*_1_ = 0.5, we have that the probability that *R*_02_takes on values less than 1 is not negligible.

**Figure 4:**
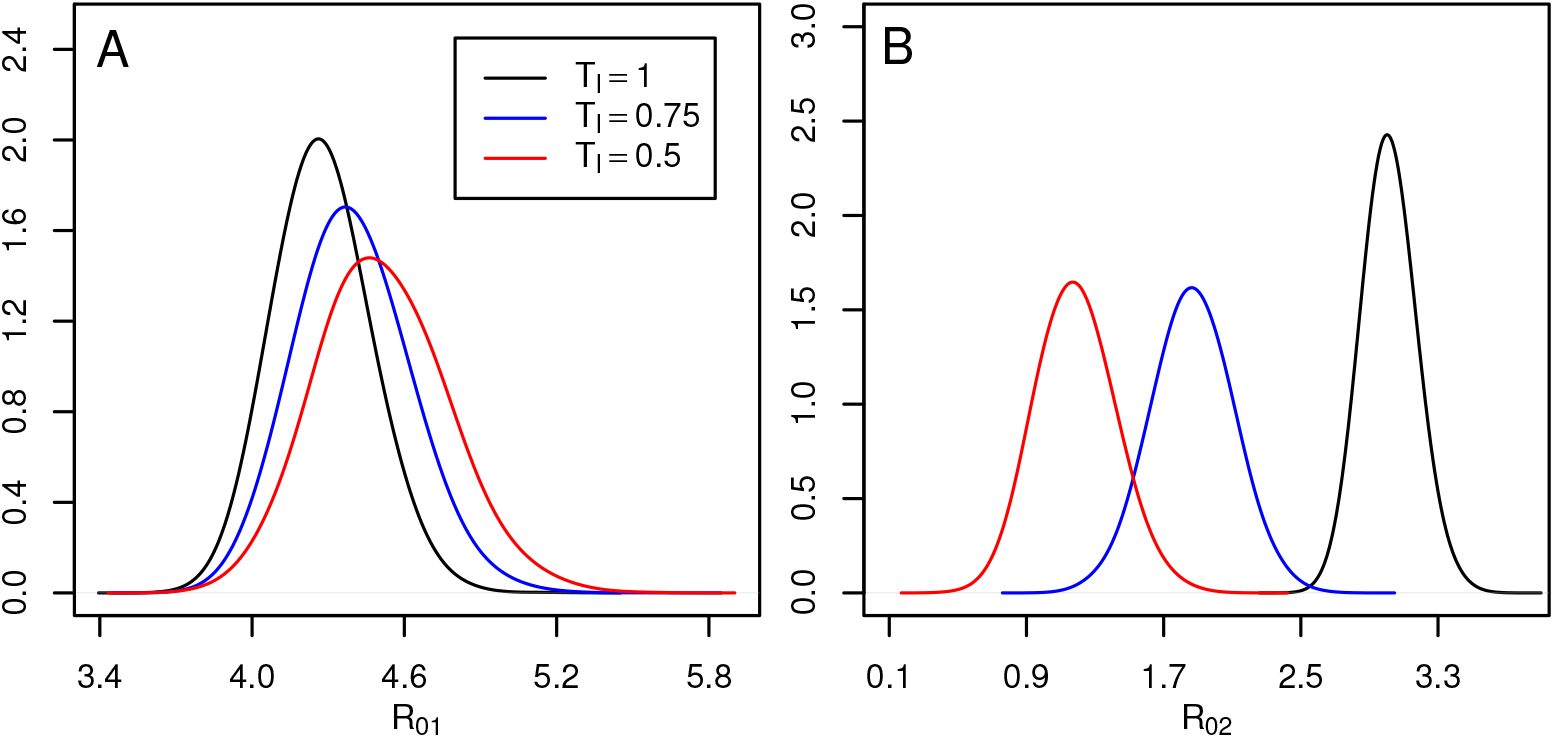
The posterior density of *R*_0_ from the north side (A) and the south side (B).

The estimates obtained above coincides with the fact that there was only 1 confirmed case of dengue on the south side from 33th to 35th week, while on the north side there were 18 cases in the same period of time, see Figure 2. Thus, the appearance of a dengue outbreak in the south region may be due to the daily movement of people. These results may suggest that not considering daily human movement overestimates the transmission rates and the local basic reproductive number of the south side of the city. The daily mobility between both patches decreased the estimated values of the local transmission rates of the South side, attributing the outbreak on that side due to the connection with the North side.

## 4. Conclusions and discussions

Many mathematical models based on ordinary differential equations have been used to model the dynamics of infectious disease throughout a country or a city. We believe it is essential to consider that an area may have, for example, different demographic and socioeconomic characteristics which may be affected by the daily mobility of people. Considering these factors for modeling the disease dynamics and estimating the model parameter are very important because takes into account heterogeneities within a region and provides a way to analyze possible effects on each local dynamics of the disease.

Here we have explored a scenario of human mobility by fitting data of a dengue outbreak from Hermosillo to a two-patch mathematical model. For the estimation process, we have only used data from the initial stage of the outbreak because there are many factors that can influence the disease dynamics for the complete outbreak. In addition, since no data are available on the mosquito population, some assumptions based on literature, were made for parameters related to the vector. On the other hand, despite not having an overall *R*_0_ of coupled model (1)-(2), the expression for the basic reproductive number given in (3) was helpful to measure the local severity of the disease in each patch.

Based on the results, we have observed that not considering the daily mobility between connected areas may lead to inappropriate conclusions of some characteristics of the disease dynamics. We have obtained higher estimates of transmission rates and local basic reproductive number in the south side if it is assumed that there is no flow of people between the north and south sides of the city. This fact could lead to suppose that the conditions for the spread of the disease on the south side may be relatively similar to the north side. However, the results may suggest that some infected residents of the south side could be a contribution of daily mobility, but not because of the conditions on that side of the city, which is consistent with previous results [1, 14, 18]. The latter could lead to more appropriate decisions regarding where to focus the control measures when an outbreak is stronger. Thus, despite having a reasonable fit of the uncoupled model to the data (case *T*_*l*_ = 1), we must take into account the importance of mobility and how this could significantly affect the dynamics in regions without conditions of disease development. Similar conclusions were obtained for other settings of the proportions that move between patches (*α*_1_ and *α*_2_, with *α*_1_ *> α*_2_).

Since human mobility has been identified as a critical factor in the spread of infectious diseases, nowadays, articles in the same line with this work are more recurrent. Furthermore, the current COVID-19 pandemic makes it even more relevant to study mathematical models with an approach that includes daily mobility. This type of study allows us to analyze the weight of commuters in the disease dynamics, which may be useful to propose control policies and reduce cases or prevent outbreaks in certain city locations.

Finally, this work is supported by the knowledge of dengue cases within a population and a socio-economic and socio-demographic analysis, to study the city as two regions connected by the human movement. However, in general, we could have a better understanding of the local properties of a community if we had more documented information about mobility. This latter could help to detect contagion risk areas within the same community.

## Data Availability

None.

